# Explainable machine learning for revisiting reported Irritable Bowel Syndrome correlates in a student cohort

**DOI:** 10.64898/2026.04.13.26350820

**Authors:** Leonardo Ramirez-Lopez, Patrick Kang

**Affiliations:** Imperial College London, Imperial College Business School, London, United Kingdom; BUCHI Labortechnik AG, Department of Data Science, Flawil, Switzerland

## Abstract

Irritable Bowel Syndrome (IBS) affects a substantial proportion of university students, yet its factors remain incompletely characterised in South Asian populations. We reanalysed a publicly available dataset of 550 Bangladeshi students from Hasan et al. [1], conducting a data audit that identified implausible records, including males reporting menstrual symptoms, and reduced the analytic sample to 506 observations. Using Explainable Boosting Machines (EBMs), which capture non-linear effects and pairwise interactions without sacrificing interpretability, we found that psychological distress, elevated BMI and academic dissatisfaction were the strongest predictors of IBS (mean AUC = 0.852 across 100 stratified train-test splits). Critically, several findings diverged from the original logistic regression analysis. Physical activity showed a non-linear risk pattern only at high intensity, the association with gender was substantially weaker when we accounted for metabolic and psychological factors as well and malnourishment does not have a strong an impact as in the original study. These divergences likely arise because the machine-learning model captures non-linear effects and interactions that were not represented in the original regression specification. Our findings underscore the value of reanalysing existing datasets with methods suited to capturing complexity and highlight data quality verification as a necessary step in the secondary analysis.

**Author summary:** We reanalysed a dataset on Irritable Bowel Syndrome (IBS) among university students in Dhaka, Bangladesh. Before modelling, we audited the dataset, removed implausible records, and reconstructed the IBS classification from the Rome III questionnaire. We then applied an interpretable machine-learning model capable of modelling non-linear effects and interactions between variables. Psychological distress (particularly anxiety and stress), body mass index, and dissatisfaction with academic major showed the strongest associations with IBS. The model also identified several interaction effects involving BMI. Our results differ in several respects from the original regression analysis, suggesting that modelling assumptions and data validation can influence the interpretation of IBS correlates. This study shows how explainable machine-learning models can complement conventional statistical analyses and how data validation can affect results in secondary analyses.

## Introduction

The gut–brain axis has been linked to conditions such as anxiety, depression, PTSD, and IBS, particularly when disruptions in the gut microbiome (dysbiosis) occur [2–4]. In this respect, psychosocial and somatic factors are increasingly acknowledged as interacting determinants of *Irritable Bowel Syndrome* (IBS), especially in young adults where these exposures often co-occur. These insights have prompted a growing interest in interventions targeting the gut microbiome, in conjunction with psychiatric approaches for the treatment of IBS [5, 6].

This study builds on recent work by Hasan et al. [1], who examined IBS prevalence among university students in Dhaka, Bangladesh, and its relationship with psychiatric and lifestyle-related factors. They reported significant correlations between IBS and psychological distress, encompassing anxiety, depression, and stress. The study aimed to estimate IBS prevalence in this population and explore how mental health and behavioural characteristics relate to the condition. However, their study appears to face four methodological limitations that, if addressed, could improve the robustness and interpretability of the findings: (*i*) substantial data anomalies potentially affecting key results; (*ii*) no examination of interaction effects, which may obscure synergistic or conditional associations with IBS. This is a common limitation in the IBS literature, which often focuses on main effects while overlooking informative interactions. Exploring interactions can enhance clinical prediction models by addressing non-additivity, improving calibration, and accounting for effect heterogeneity [7]; (*iii*) discretisation of continuous variables (e.g. psychiatric scores, BMI), which aids interpretation but reduces power and may mask non-linear associations; and (*iv*) the assumption of linearity between IBS and explanatory variables, which can yield biased estimates when associations are non-linear. These limitations are not only technical, but we demonstrate that addressing this leads to substantively different conclusions regarding the predictors of IBS in this population, including the role of BMI, physical activity and gender.

The present study has two objectives. First, we perform a methodological audit of the dataset and an analytical strategy used by Hasan et al. [1], identifying inconsistencies in the data and modelling choices that may affect the validity of the reported associations. Second, we reconstruct the analytical dataset and conduct a reanalysis using an interpretable machine learning framework capable of modelling non-linear effects and interactions between psychiatric and somatic variables and incorporating possible interaction effects. While causal inference is beyond the scope of this analysis, our aim is to reconcile predictive accuracy with interpretability, two aspects often treated as a trade-off. By using Explainable Boosting Machines, we offer a framework that balances predictive performance with transparency, allowing domain experts to evaluate and refine mechanistic hypotheses about IBS without sacrificing interpretability for accuracy.

## Materials and methods

### Data source

The dataset analysed in this study derives from a cross-sectional questionnaire survey of undergraduates in Dhaka, Bangladesh, originally conducted by Hasan et al. [1]. Thus, no new human participants were recruited and no additional data were collected for the purposes of this study. We used the original data provided in Hasan et al. [1] which was accessed and retained for the present study on 14 July 2025 (see S1 Appendix). For this study, we did not have access to information that could identify individual participants. The original study employed validated instruments, including the Rome III criteria for IBS and the DASS-21 scale for psychological distress. The survey captured a broad range of sociodemographic, psychological, and lifestyle factors.

### Data validation and reconstruction

The dataset analysed in this study was obtained from the original survey published by Hasan et al. [1]. It includes variables from multiple domains relevant to IBS risk, including sociodemographic characteristics (e.g., age, gender, residence type), psychological health indicators (depression, anxiety, and stress measured using DASS-21), lifestyle behaviours (e.g., physical activity, diet, smoking, alcohol consumption), academic satisfaction variables, and anthropometric measures (height, weight, and body mass index, BMI). The original dataset contained 550 observations. A detailed data dictionary describing all variables and coding schemes is provided in S5 Appendix.

Prior to modelling, the dataset was subjected to a validation and preprocessing procedure to identify anomalous observations and ensure consistency in the construction of the outcome variable. Several inconsistencies were detected in the raw data obtained from the original study [1], which required filtering and reconstruction steps before analysis.

First, we used anthropometric plausibility filters to remove physiologically unrealistic measurements. Four records were excluded based on the following criteria: height greater than 300 cm, weight less than 0 kg or greater than 200 kg, and body mass index (BMI) values below 10 or above 40.

Second, the IBS outcome variable was reconstructed from the responses to the Rome III diagnostic items. During this process, we detected a data integrity issue in which 40 respondents classified as male reported menstrual-related abdominal discomfort. Given the biological inconsistency between these responses and the reported gender, these observations were excluded from the analysis. Note that this filtering step is not reported to have been undertaken in the original publication by Hasan et al. [1] which may affect the validity of their reported estimates.

After applying these validation and reconstruction procedures, the final analytic sample consisted of 506 students.

### Outcome variable

The objective of the modelling framework was to estimate the association between Irritable Bowel Syndrome (IBS) and a set of demographic, behavioural, anthropometric, academic, and psychological variables. The response variable was a binary indicator representing the presence of IBS based on the reconstructed Rome III diagnostic criteria. Let *Y*_*i*_ denote the IBS status of individual *i*, where

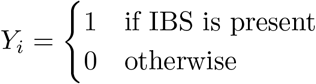

Predictors included sociodemographic characteristics (e.g. gender, age group, education level, marital status), academic and financial variables (e.g. major choice, satisfaction with major, funding source), lifestyle behaviours (e.g. smoking, physical activity, breakfast behaviour, diet, sleep duration), anthropometric measures (BMI), and psychological scores derived from the DASS-21 instrument (depression, anxiety, and stress). A detailed description of all variables and coding schemes is provided in S5 Appendix.

### Explanatory variables

Explanatory variables were selected based on theoretical relevance to IBS and consideration of potential multicollinearity. Variables corresponding to the Rome III diagnostic criteria were excluded from the explanatory set because they were used to reconstruct the IBS outcome variable. Including these items as predictors would result in circular prediction and artificially inflated model performance.

Psychological variables were derived from the Depression Anxiety Stress Scales (DASS-21). The individual questionnaire items were not included as explanatory variables because they are aggregated into the depression, anxiety, and stress subscale scores. Including both item-level responses and composite scores would introduce substantial multicollinearity without providing additional information. Binary indicators derived from these scores were also excluded because they represent categorised versions of the continuous measures and contain less information. The continuous DASS-21 scores were used directly in the modelling framework, while categorical severity thresholds were applied only for descriptive summaries.

Anthropometric information was represented using body mass index (BMI). Height and weight were not included separately as explanatory variables because they are combined into BMI, which is a standard clinical indicator of body composition.

The final set of explanatory variables comprised demographic characteristics (gender, age group, education level, marital status), academic and financial factors (choice of major, satisfaction with major, funding source), lifestyle behaviours (smoking, physical activity, breakfast habits, fruit and vegetable consumption, sleep duration), BMI, and continuous psychological scores for depression, anxiety, and stress.

### Modelling framework

In general terms, the modelling objective can be expressed as

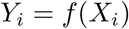

where *X*_*i*_ represents the vector of explanatory variables for individual *i* and *f* (·) denotes an unknown function relating these predictors to the probability of IBS. To estimate this relationship, we employed Explainable Boosting Machines (EBMs) [8]. EBMs belong to a class of generalised additive models trained using boosting. It combines the predictive performance of modern machine learning methods with the interpretability of additive statistical models. This modelling approach was selected for two main reasons. First, EBMs can reliably capture non-linear relationships between predictors and the outcome without needing to explicitly specify the functional forms. This is particularly relevant in the context of IBS, where variables such as BMI or psychological distress may exhibit non-linear associations with disease risk. Second, EBMs retain an interpretable structure through additive component functions. This allows the visualisation of the marginal effect of each predictor. In addition, EBMs can estimate pairwise interaction terms, and enable the exploration of potential synergistic relationships between variables.

In contrast to the original study, which relied on conventional regression modelling and discretised several continuous predictors, we trained an EBM using the `ebm` package [9] in `R`. The model used the demographic, lifestyle, anthropometric, and psychological variables described above to predict IBS presence.

As 118 individuals were classified as having IBS and 388 as not having IBS, the dataset exhibited a clear class imbalance. For that, we applied random oversampling of the minority class within each cross-validation split. Psychological scores were retained as continuous variables and were subject to monotonic constraints based on prior hypotheses. Model performance was evaluated across 100 stratified train (75%) and test (25%) splits. We report mean accuracy, sensitivity, specificity, F1 score, and AUC across replications.

The final model was trained on the full balanced dataset using log-loss as the optimisation objective. Model interpretation relied on the additive and pairwise shape functions produced by the EBM, which quantify the marginal or interaction contribution of each predictor to the predicted log-odds of IBS.

## Results

### Exploratory analyses

The distributions of variables in our sample differ slightly from those reported by Hasan et al [1] because 44 observations with implausible or inconsistent values were excluded according to the data cleansing criteria described in the previous section.

The gender distribution in the sample of students is approximately balanced, with 49.6% as male and 50.4% female. This near-equal split supports gender comparability in subsequent analyses. Nearly half of the students (49%) were between 19 and 24 years of age. Over half of the students (55%) reported skipping breakfast on three or more days per week, while 45% skipped breakfast less frequently. Additionally, 21% of participants identified as smokers. Approximately 37% of students engaged in less than 20 minutes of physical activity per day, a pattern consistent with sedentary behaviour profiles reported among university students in this sample.

Table 1 presents the distribution of selected categorical variables, along with the percentage of participants within each group reporting IBS, elevated depression, elevated anxiety and stress symptoms. The thresholds for depression and anxiety were derived from the DASS-21 scoring guidelines [10], and elevated symptoms were defined as scores ≥ 10 for depression, ≥ 8 for anxiety, and ≥ 13 for stress corresponding to severe or higher severity levels. These cutoffs are commonly used in psychological research to identify clinically meaningful levels of emotional distress.

**Table 1.**
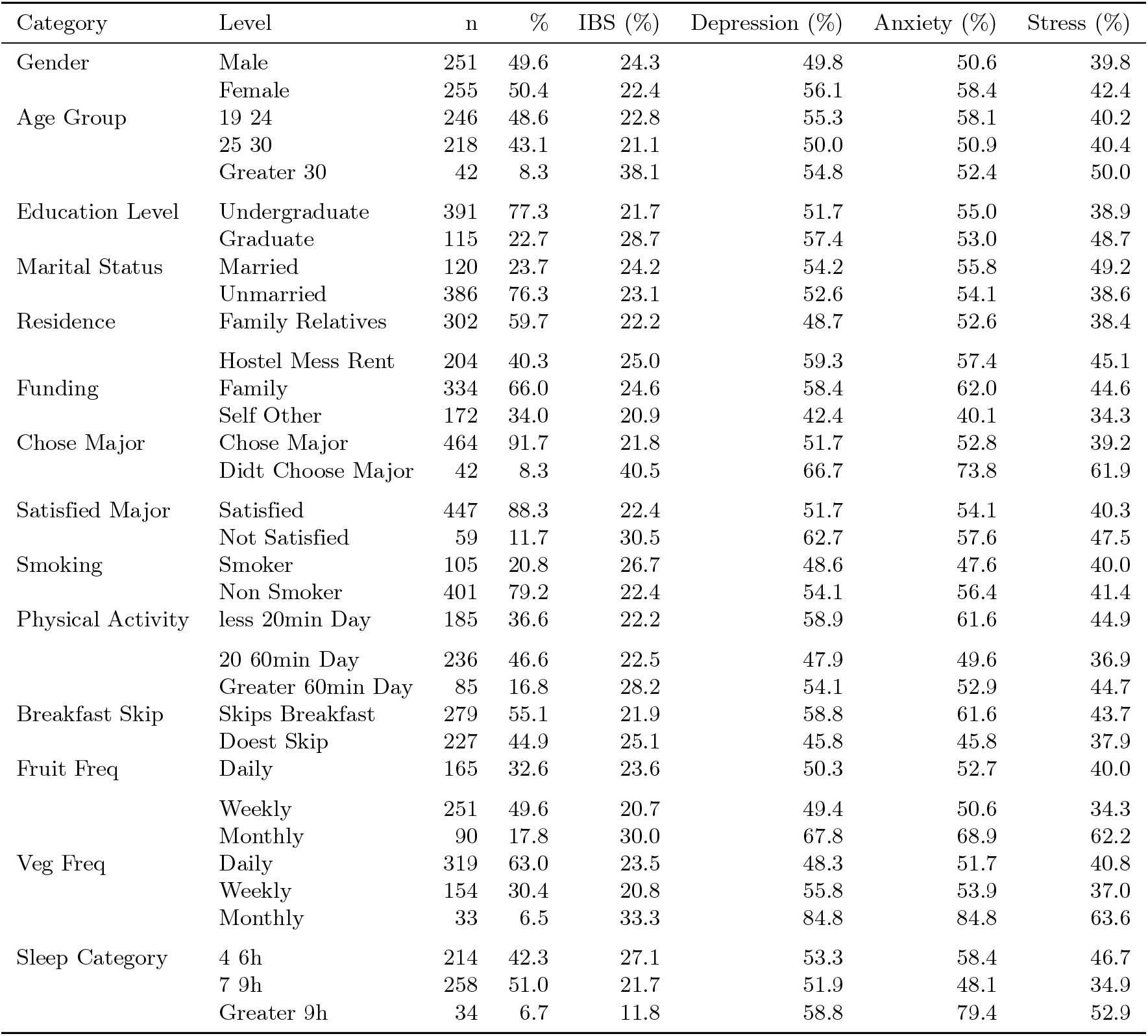
Summary of categorical variables by IBS, depression, and anxiety status.

Based on the application of Rome III diagnostic criteria to the available questionnaire data, the prevalence of IBS in the sample was approximately 23.32 %. This estimate is lower than originally reported by Hasan et al. [1], yet it exceeds national community-based prevalence rates in Bangladesh (6.5–7.8%) [11].

For descriptive summaries, DASS-21 subscale scores were categorised using standard severity thresholds, whereas continuous scores were used in the modelling framework. Mean scores were 11.2 (sd = 10.2) for depression and 10.0 (sd = 8.7) for anxiety, indicating severe and extremely severe levels. Violin plots (Fig 1) show higher medians and greater variability among IBS students, especially for depression, with the IQR shifted upward. This highlights a disproportionate psychological burden. These findings support the use of monotonic constraints in EBM, ensuring risk increases with higher scores, and align with prior studies linking IBS to elevated psychological distress [12, 13].

**Fig 1.**
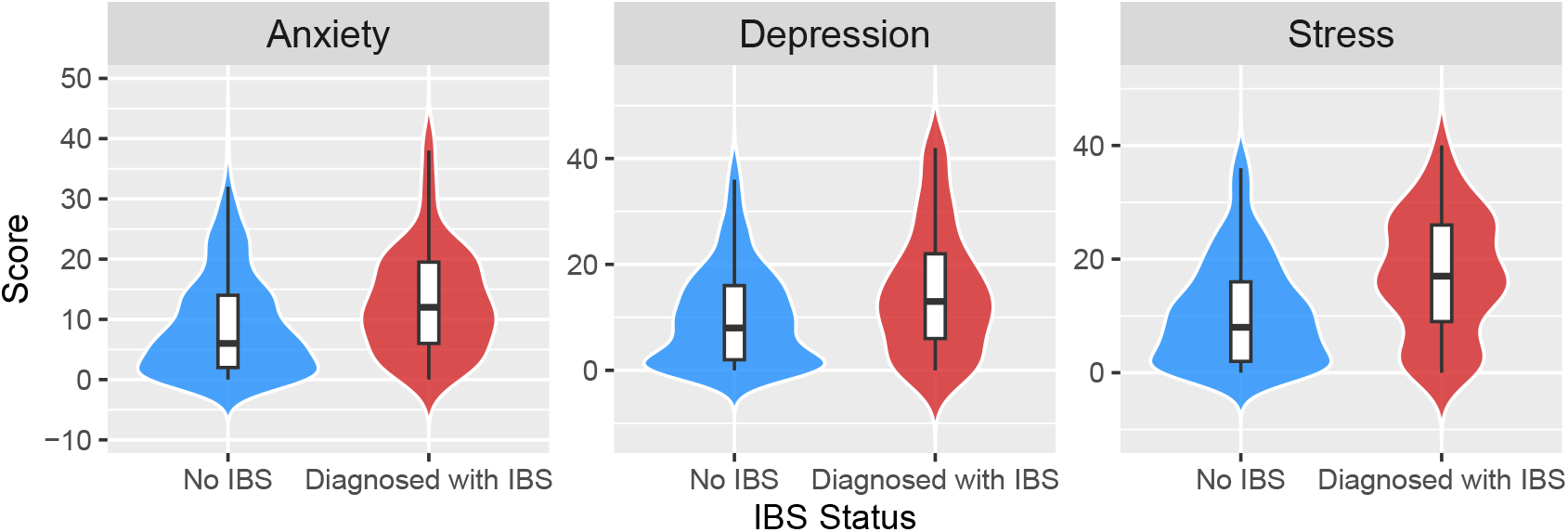
Distribution of depression and anxiety scores by IBS status.

### Model-derived associations with IBS

Our EBM model achieved an average AUC of 0.852 (sd = 0.055) across 100 random replications, which indicates good predictive performance. The F1 score averaged 0.911 (sd = 0.026), with sensitivity and specificity of 0.91 (sd = 0.036) and 0.716 (sd = 0.093), respectively. In this respect, our most important performance figure is sensitivity, which reflects the ability of the model to correctly identify individuals with IBS. As for ROC, our results outperform those originally reported by Hasan et al. [1] where AUC was 0.7486. Looking at these performance metrics, the EBM model appears to capture consistent statistical associations between the explanatory variables and IBS.

Fig 2 shows model variables ranked by their mean absolute contribution to the predicted log-odds of IBS presence, averaged across all observations and bagged estimators in the final EBM model. This score reflects the overall influence of the explanatory variables on predictions. To enhance interpretability, we applied a cumulative importance threshold. This retains only features that together explained 70% of the total importance. We focused our interpretation on the first 13 features in the ranking (see Figure 2), these collectively accounted for over 70% of the cumulative importance. They include both main effects (e.g. `bmi_sc`, `anx_score`, `str_score`) and a subset of interaction terms. Of the 16 interactions considered by the EBM model, only 4 surpassed the cumulative importance threshold, suggesting limited but specific synergistic effects. In contrast, some variables often considered relevant (e.g. `gender`) ranked among the least association features. The two variables with the lowest contribution were `education_level` and the interaction `age_group & physical_activity`. Although our model does not apply explicit regularisation, the application of L1 or L2 penalties could help further refine feature selection.

**Fig 2.**
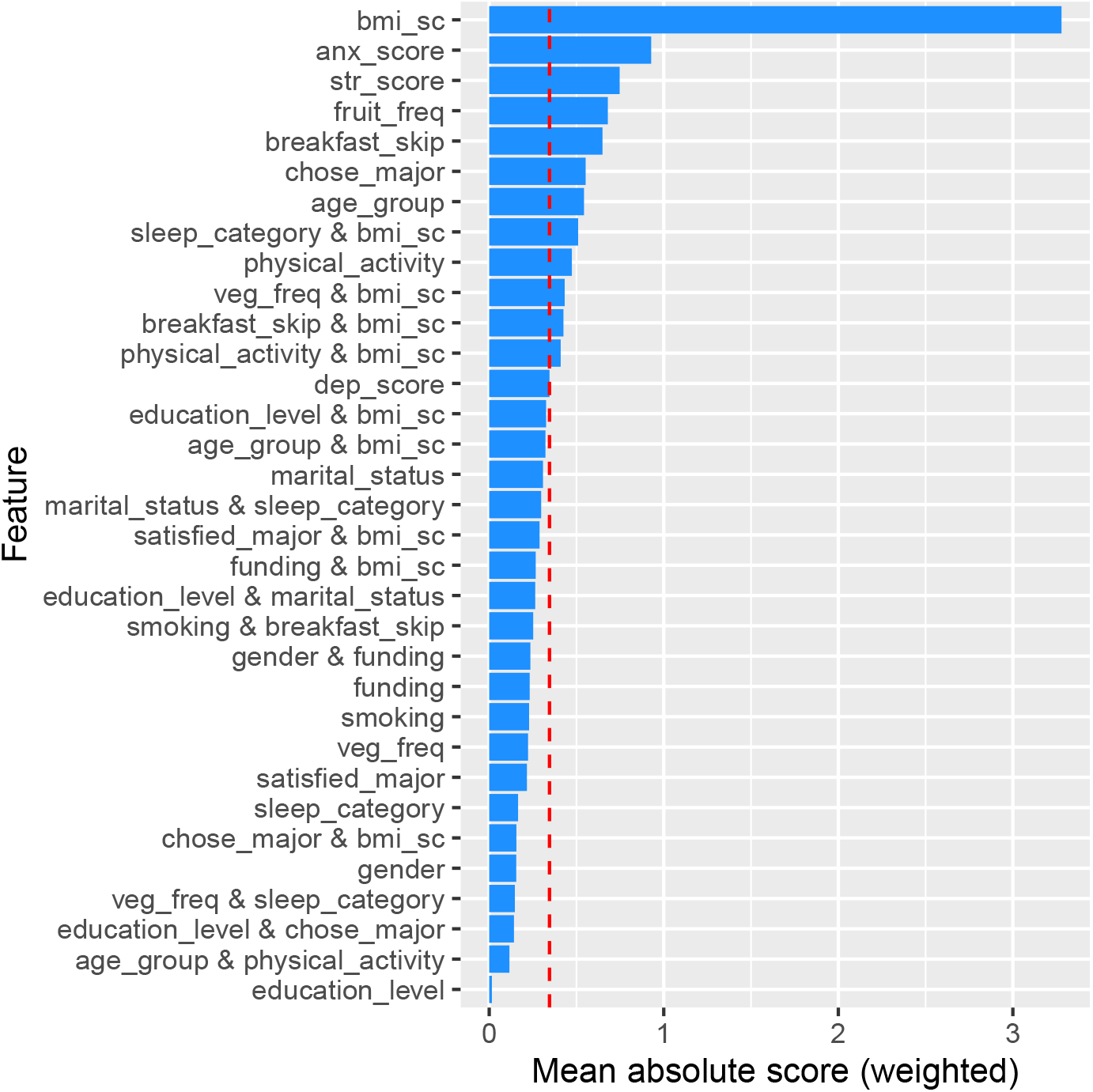
Ranking of the contribution of the variables and interactions in the EBM model. The red line indicates the threshold for cumulative importance.

Fig 3 shows the additive contributions of continuous predictors of the log-odds of IBS in the EBM model. Positive values indicate an increased likelihood of IBS. Shaded ribbons represent 95% confidence intervals, indicating overall stability of the estimates. For BMI (`bmi_sc`), the contribution seems to be random across the lower range but shows a consistent increase beyond a BMI of 30, suggesting non-linearity and also an elevated IBS risk in individuals with obesity. For anxiety (`anx_score`), stress (`str_score`), and depression (`dep_score`), the risk of IBS increases with symptom severity. This trend reflects domain-informed monotonicity constraints imposed during model training. Marked increases in log-odds at high scores highlight the disproportionate impact of severe psychological symptoms on IBS risk.

**Fig 3.**
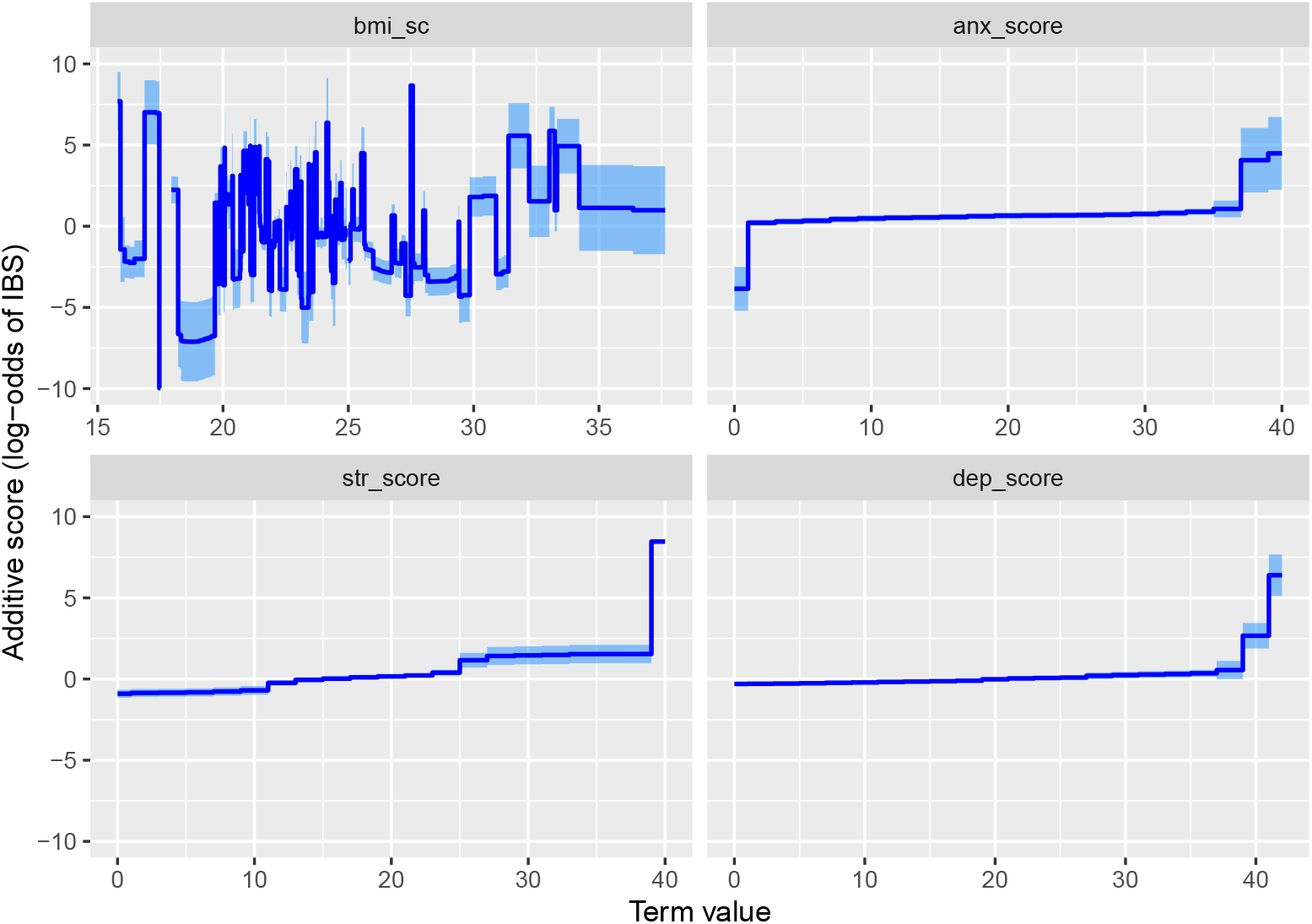
Additive contributions of continuous predictors to IBS log-odds in the EBM model.

For our categorical variables, Fig 4 shows their additive contributions to IBS. Monthly fruit consumption is associated with elevated IBS risk, while daily and weekly intake have negligible effects. Skipping breakfast lowers IBS risk relative to regular breakfast consumption. Not choosing one’s academic major is strongly linked to increased IBS likelihood. Individuals aged over 30 show the highest risk; those aged 25–30 exhibit reduced risk, and the 19–24 group shows no significant effect. Physical activity shows a non-linear pattern: low to moderate activity reduces IBS risk, while high levels (*>*60 minutes/day) increase it. Note that the bars (which indicate 95% confidence intervals), reflect the uncertainty around the estimated effects; the largest uncertainty is observed for the `did not choose major` and `age` *>*`30` categories.

**Fig 4.**
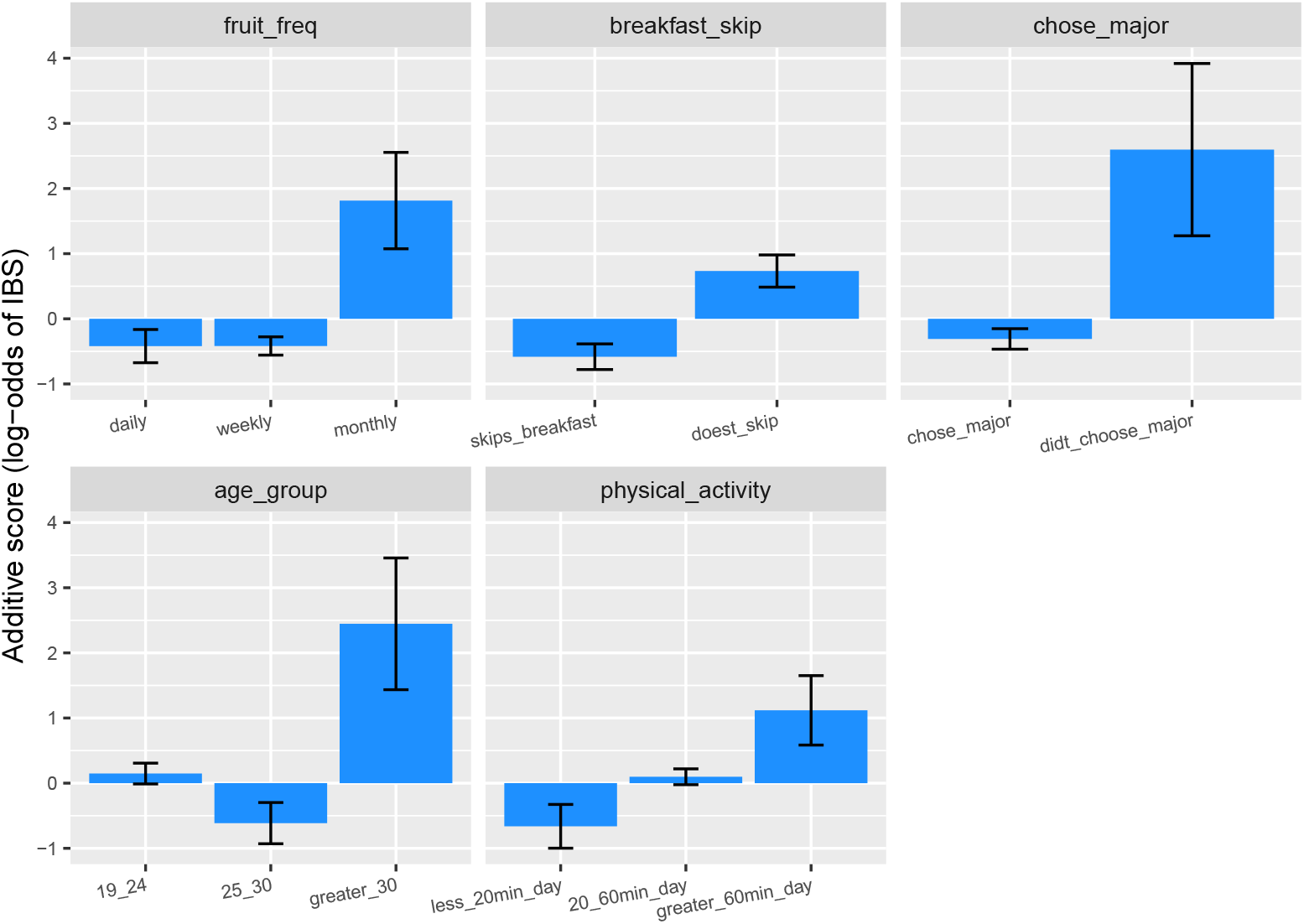
Additive contributions of categorical predictors to IBS log-odds in the EBM model.

Figure 5 displays the estimated interaction effects between BMI and selected categorical predictors on IBS log-odds. These interactions are largely driven by BMI, whose main effect was stable only for values above 30 (i.e. in the obesity range). Accordingly, interpretations focus on this range. For the `bmi_sc` × `breakfast_skip` interaction, IBS risk is the highest among individuals with elevated BMI who do not skip breakfast. In the `bmi_sc` × physical activity panel, IBS risk increases with BMI and physical activity. This suggests a potential non-linear modulation of IBS risk in obese individuals who engage in high physical activity. The `bmi_sc` × `sleep_category` interaction is less clearly defined, but hints towards a general increase in IBS risk with rising BMI. A slight reduction in risk is observed with longer sleep durations. Similarly, in the `bmi_sc` × `veg_freq` interaction, IBS risk increases with BMI, with a modest attenuation among those reporting more frequent vegetable consumption.

**Fig 5.**
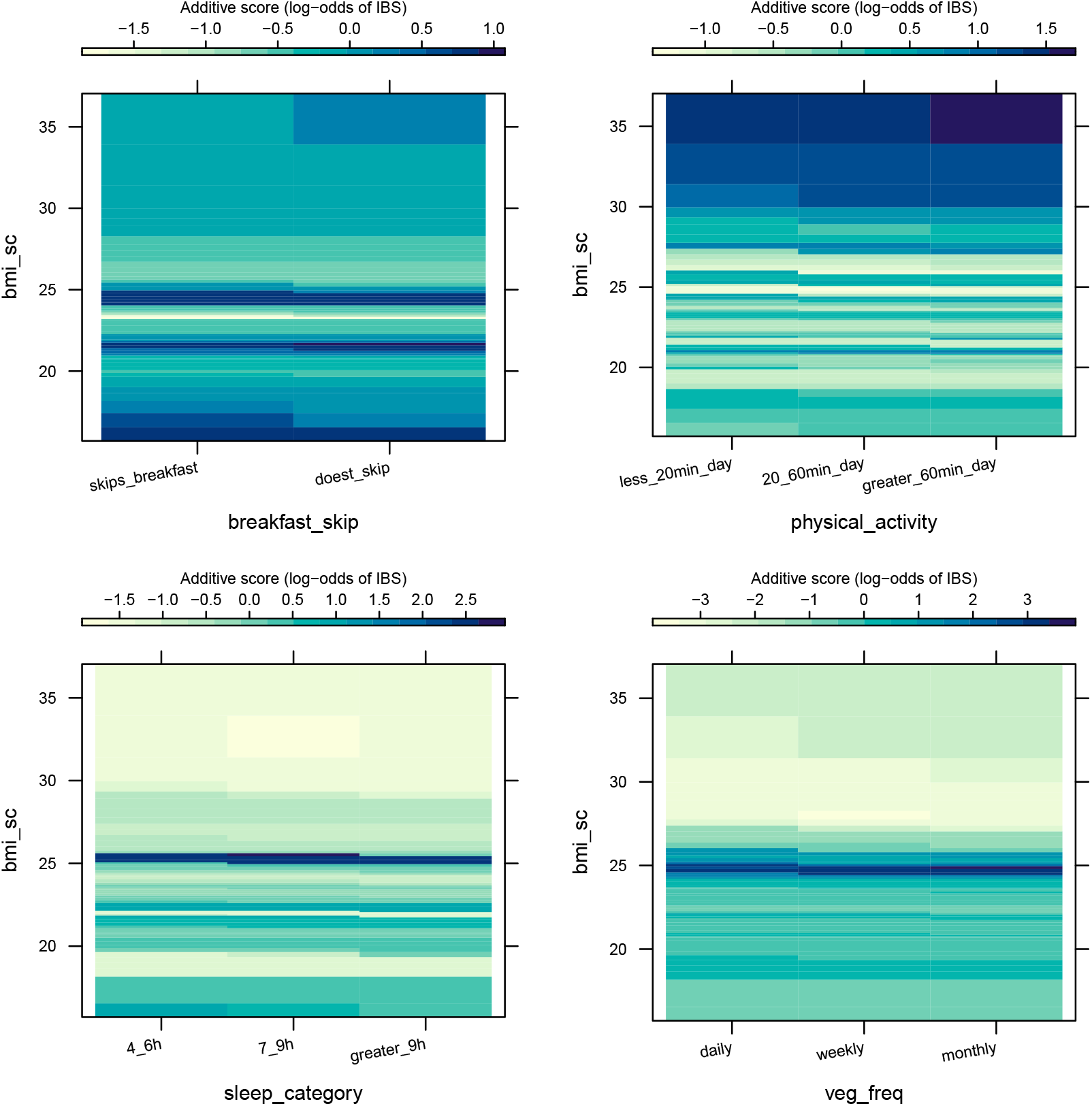
Additive contributions of interactions to IBS log-odds in the EBM model.

## Discussion

Our reanalysis of the Hasan et al. [1] dataset yielded two distinct contributions. First, a data audit identified systematic anomalies in the source data, including males reporting menstrual symptoms, which were not addressed in the original analysis and could have introduced misclassification bias in the reported association. Second, applying Explainable Boosting Machines to the cleaned dataset achieved substantially higher predictive accuracy (AUC = 0.852 compared to 0.7486) and revealed several associations that diverged from the original findings coming from the logistic regression. These divergences likely arise because the machine-learning model captures non-linear effects and interactions that were not represented in the original regression specification, particularly those involving BMI as a moderating variable.

Despite these divergences, our findings converge with those of Hasan et al. [1] on the central role of psychological distress in IBS. Anxiety and stress emerged as the strongest and most consistent predictors in both analyses, while depression showed a comparatively weaker association in each. This agreement across methodologically differing approaches strengthens confidence that psychological distress is a robust correlate of IBS in this population, independent of modelling assumptions. The gut–brain axis literature provides a plausible basis for these associations, with dysbiosis and neuroendocrine dysregulation proposed as the mechanism. [2–4].

Despite methodological differences, our analyses and those of Hasan et al. [1] show agreement on only a limited number of findings. In both studies, anxiety and stress emerged as important variables associated with IBS, whereas depression showed a comparatively weaker association. Beyond these psychological factors, however, the overall patterns differ substantially. Several lifestyle, demographic, and behavioural predictors identified in the original analysis either show different effect structures or reduced importance in the machine-learning model, leading to contrasting interpretations of the factors associated with IBS in this population.

Unlike the original study, which linked IBS risk to malnourishment, our findings point to high BMI (especially in the obese range) as a stronger predictor. While breakfast skipping did not show a significant association with IBS in the original study, our analysis indicates that this behaviour is associated with a lower estimated risk of IBS (Fig 4). The interaction effect suggest that this association is not uniform across BMI categories, with the protective pattern most pronounced among individuals with elevated BMI. Another possible interpretation is that this pattern may reflect differences in meal timing or eating regularity, although this hypothesis remains speculative and warrants further investigation [14].

Another contrasting finding concerns gender. While female gender emerged as a significant predictor in the original regression analysis, it showed low predictive importance in the machine-learning model. This is consistent with how tree-based models handle correlated predictors. When stronger variables such as psychological distress and BMI are present, demographic variables that partially overlaps with them tend to lose their independent importance. This does not imply that gender is clinically irrelevant but rather that its association with IBS in this sample may be mediated through psychological and metabolic pathways rather than operating as an independent risk factor.

Hasan et al. [1] suggest that higher levels of physical activity may have a protective association with IBS. Our findings do not indicate that regular physical activity is detrimental, however, they reveal a non-linear relationship, with increased IBS risk observed at very high activity levels (*>* 60 minutes per day; Fig 4). Several studies have suggested that intense activity levels may elevate the risk of gastrointestinal disturbances by inducing physiological changes such as reduced intestinal blood flow, epithelial injury, impaired barrier function, altered gut motility, and inflammatory responses [15–17] This non-linear pattern was further moderated by BMI, with elevated risk concentrated among individuals in the obese range. This suggests that physical vulnerability could amplify the gastrointestinal stress associated with intense exercise.

In contrast to the original analysis, which suggested a general association between fruit consumption and IBS, our model indicates that the relationship is not monotonic (Fig 4). Only infrequent fruit intake (monthly consumption) was associated with elevated IBS risk, whereas weekly or daily consumption showed negligible effects. This suggests that the association may reflect broader dietary patterns rather than a direct effect of fruit intake. Some studies report lower fruit and vegetable intake among individuals with IBS and link poorer diet quality with more severe symptoms [18]. However, it is important to note that some fruits can trigger symptoms due to FODMAP sugars (fructose, sorbitol).

We also found that respondents who reported not having chosen their academic major had a higher predicted risk of IBS, potentially reflecting greater psychological distress, consistent with known IBS–mental health associations. Older participants (*>*30 years) exhibited elevated IBS risk, possibly due to cumulative stress or comorbidities (Fig 4). High physical activity also predicted IBS, which, while unexpected, may reflect symptom exacerbation in subgroups with vulnerabilities such as obesity. These patterns highlight the moderating role of BMI and underscore non-additive interactions between physiological and behavioural factors.

These findings should be interpreted in light of several limitations. Although 44 anomalous observations were removed, additional inconsistencies may remain in the source dataset. The cross-sectional design precludes causal inference, and the restricted sample may limit generalisability. Institutional clustering was not modelled and may affect uncertainty estimates. Nevertheless, the interpretable machine-learning framework enabled the modelling of non-linear effects and interactions among predictors, revealing patterns (particularly involving BMI) that were not captured in the original regression analysis.

## Conclusion

This study revisited the correlates of *Irritable Bowel Syndrome* (IBS) in university students using an interpretable machine-learning framework capable of incorporating domain knowledge, modelling non-linear effects, and capturing interactions among predictors. The analysis highlights psychiatric and behavioural variables (particularly anxiety, stress, higher BMI, and dissatisfaction with the chosen academic major) as key contributors to IBS risk. The inclusion of interaction terms further revealed non-additive relationships among predictors, notably those involving BMI.

Compared with the original regression-based analysis reported by Hasan et al. [1], several predictors exhibited different importance or effect structures, particularly the roles of BMI, physical activity, and gender, indicating that interpretation of IBS correlates in this dataset depends strongly on modelling assumptions and data pre-processing.

The modelling framework demonstrates that predictive performance and interpretability can be jointly achieved, providing a transparent alternative to conventional statistical approaches. Such approaches are particularly valuable in epidemiological research, where the objective is not only prediction but also the generation of plausible mechanistic hypotheses. At the same time, the reliability of insights derived from this dataset depends critically on data quality. Several biologically or logically inconsistent responses were identified, emphasising the importance of rigorous data validation when reusing publicly available datasets. Clarification of these anomalies would help ensure the reliability of future secondary analyses.

## Data Availability

The data used in this study originate from the publicly reported dataset described by Hasan et al. All data necessary to reproduce the analyses in this study, together with the code and detailed methodological steps used for data processing and modelling, will be made publicly available in an open repository upon acceptance of the manuscript.

## Supporting information

**S1 Appendix. Link to the original data**. The original data produced by Hasan et al. [1], was obtained from the following link: https://doi.org/10.1371/journal.pgph.0004670.s001. The data used in the present study was accessed and retained for the present study on 14 July 2025.

**S2 Appendix. Link to the original ROME III questionnaire for IBS diagnostics**. The original questionnaire employed by Hasan et al. [1], can be obtained from the following link: https://doi.org/10.1371/journal.pgph.0004670.s002

**S3 Appendix. Cleaned and reconstructed dataset**. The cleaned dataset and the reconstructed IBS classification used in this study will be made publicly available in an online repository upon acceptance of this manuscript. A permanent access link will be provided here.

**S4 Appendix. Code**. The code used for data processing, modelling, and analysis in this study will be made publicly available in an online repository upon acceptance of this manuscript. A permanent access link will be provided here.

**S5 Appendix. Data dictionary specific to this study**.

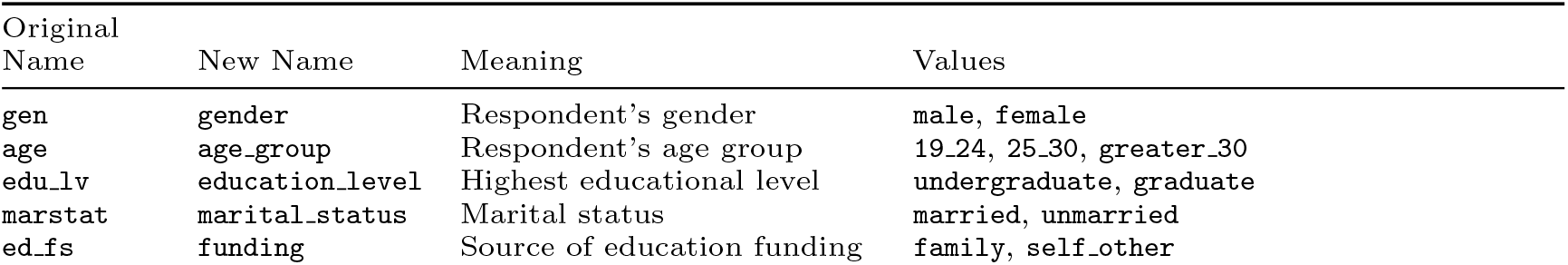
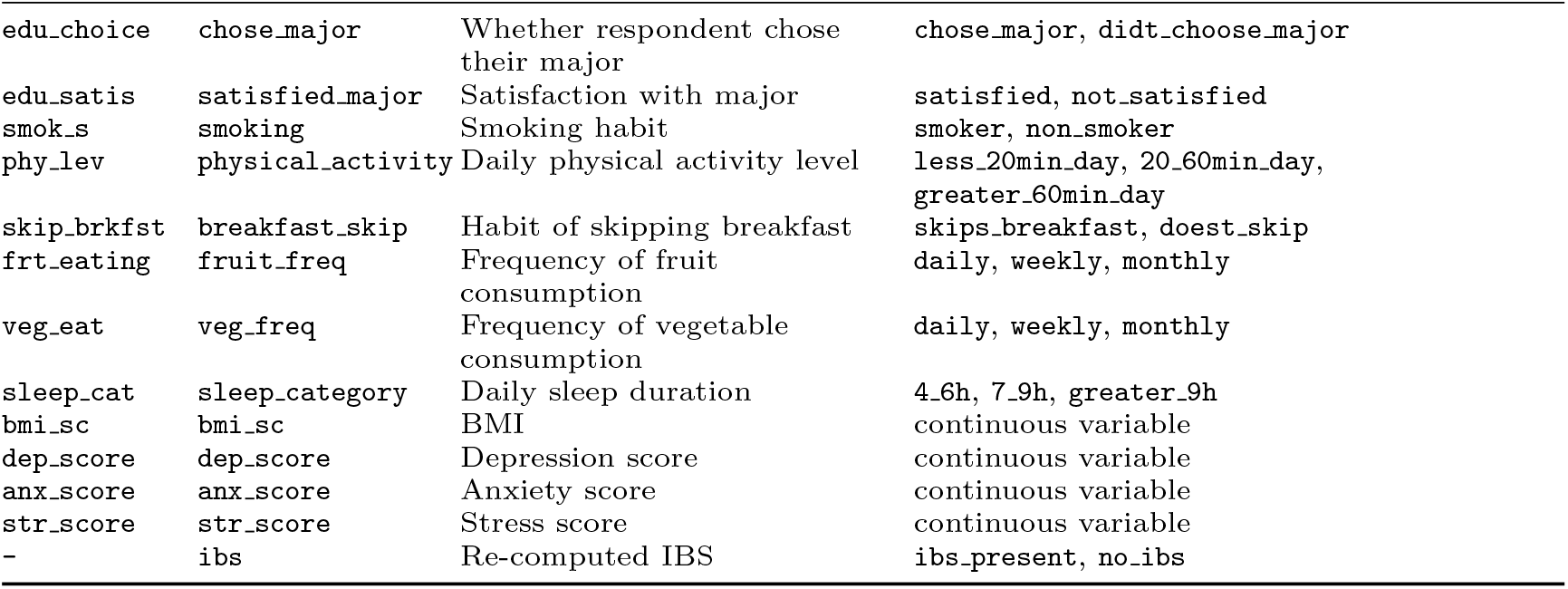

## Acknowledgments

We thank Dr Laure de Preux (Imperial College London) for her guidance and feedback on early versions of this manuscript. We also thank our colleagues Salma Abubakr, Maria Linardou, and Simon Sumer for insightful discussions that contributed to the early development of the ideas presented in this work.

